# Comparison of IA-2 Bridge ELISA and Radiobinding Assays for Progression Risk Assessment in Early-Stage Type 1 Diabetes

**DOI:** 10.64898/2026.01.26.26344598

**Authors:** Ezio Bonifacio, Marlon Scholz, Andreas Weiss, Anette-Gabriele Ziegler

## Abstract

Stratifying progression from early-stage type 1 diabetes to clinical disease is essential for optimally timing disease-modifying therapies. We previously developed a progression likelihood score (PLS) that includes quantitative IA-2 autoantibody (IA-2A) measurements. This study aligned IA-2A thresholds used for PLS calculation between the radiobinding assay (RBA) and a commercially available RSR IA-2A ELISA to support broader clinical application. Serum samples from 349 children with stage 1 type 1 diabetes were analyzed using both assays. IA-2A positivity was similar by RBA (61.6%) and ELISA (59.0%). Centile-based alignment of ELISA-positive samples defined thresholds corresponding to RBA IA-2A categories. ELISA-derived PLS low (PLS < 0.5), moderate (PLS 0.5-4.0) and high (PLS > 4.0) risk groups stratified progression to stage 3 disease comparably to RBA-derived groups. The 3-year progression rate for children with an ELISA IA-2A PLS >4.0 was 52.4% (95% CI, 30.5– 66.1), similar to the RBA-derived PLS >4.0 group (58.7%; 95% CI, 37.1–72.8). These results demonstrate that the commercial ELISA can be used for PLS-based risk stratification.

## Background

Type 1 diabetes can be diagnosed in early presymptomatic stages defined by two or more islet autoantibodies without (Stage 1) and with accompanying dysglycemia (Stage 2)^1^. Stage 2 disease is associated with a relatively rapid progression to stage 3^2^, providing opportunities to assess the efficacy of potential disease-modifying therapies to delay the onset of stage 3. These individuals, however, represent a minority of those diagnosed with early-stage disease, which can lead to long recruitment periods for clinical trials. We have developed a Progression Likelihood Score (PLS), which stratifies progression rates in children with stage 1 disease and identifies a significant number with a PLS >4.0 in whom progression rates were similar to those seen in Stage 2 disease^3, 4^. The PLS includes quantitative measurement of IA-2 autoantibodies, which are a marker for faster progression^5^. IA-2 autoantibodies were measured by harmonized radiobinding assays^6^, which are not certified assays and are not readily available or used in clinical practice, thereby limiting the use of PLS for eligibility in clinical trials. Assays often used by clinical laboratories are a bridge ELISA produced by RSR limited and commercialised as the RSR IA-2 Autoantibody ELISA version 2 or as the KRONUS IA-2 autoantibody (IA-2Ab) ELISA kit. The objective of this study was to align the thresholds required to calculate the PLS across the IA-2 RBA and commercially available ELISA assays.

## Methods

Serum samples from 349 children with stage 1 type 1 diabetes participating in the Bavarian Fr1da study were used for this comparison^4, 7^. Samples were taken at the same visit at which an oral glucose tolerance test (OGTT) and HbA1c assessment were performed; the 90-minute glucose during OGTT, the HbA1c, and the IA-2A category defined by IA-2A titre were used to calculate the PLS^3^. Stage 1 was defined as normal glucose tolerance—fasting plasma glucose <5.6 mmol/l (100 mg/dl), 2-h OGTT plasma glucose <7.8 mmol/l (140 mg/dl), and OGTT plasma glucose <11.1 mmol/l (200 mg/dl) at 30, 60, and 90 minutes—together with HbA1c <39 mmol/mol (5.7%). Children were followed for the development of stage 3, which was defined using the 2025 ADA criteria^8^.

IA-2A were measured by RBA as previously described^6^ and using the RSR IA-2 Autoantibody ELISA version 2 assay (RSR Ltd, Cardiff UK) following the manufacturer’s instructions. The threshold for positivity defined by the manufacturer was 7.5 U/ml and the range of measurement on undiluted serum was 0.95 U/ml to 4,000 U/ml. The PLS was calculated using the formula: exp[(HbA1c[%] − 5.233) × 1.125 + (OGTT90[mg/dL] − 107.6) × 0.0195 + (IA-A *cat* − 1.27) × 0.662)]^3, 4^. For the RBA measures, the IA-2A categories were previously defined as <3 (negative, category 0), >3 – 100, (category 1) >100 – 290 (category 2), and >290 (category 3). For the PLS, three risk groups were previously defined as low (PLS < 0.5), moderate (PLS 0.5 – 4.0), and high (PLS >4.0). Progression to stage 3 was assessed for each risk group using the Kaplan-Meier method with follow-up from the date of the measured PLS until the date of onset of stage 3 or the last contact if the participant had not developed stage 3.

## Results

Of the 349 samples measured by both RBA and ELISA, a positive value for IA-2A was observed in 215 (61.6%) when measured by RBA and in 206 (59.0%) when measured by the ELISA (*P*=0.54), including 193 (55.3%) positive in both assays (Figure 1). Among those who were RBA positive, the frequencies in each of the PLS IA-2A categories were 31% (66/215) for category 1, 26% (57/215) for category 2 and 43% (92/215) for category 3. To align thresholds between the RBA and the ELISA, the corresponding centiles were examined in the ELISA positive samples and corresponded to 7.5-114 units for category 1 (61/205, 30%), 115-1340 (54/206, 26%) for category 2, and >1340 for category 3 (91/206, 44%). Using these thresholds, a PLS based on IA-2A ELISA values was calculated for 347 children. Of these, 310 (89%) were assigned to PLS risk groups that were the same as the RBA IA-2A–derived PLS groups, 24 (7%) to a higher risk group, and 13 (4%) to a lower risk group than those derived from the RBA IA-2A PLS (Figure 1b). The frequency of children in the low, moderate and high risk groups was similar using the RBA and ELISA IA-2A derived PLS risk groups (Figure 1c). In particular, the high risk group was assigned to 38 children by the RBA IA-2A-derived PLS and 43 by the ELISA IA-2A-derived PLS. Furthermore, the ELISA IA-2A derived PLS risk groups were able to stratify progression to stage 3 in a similar manner to the RBA IA-2A derived PLS groups (Figure 1d). The 3-year progression rate in those with an ELISA IA-2A PLS >4.0 was 52.4% (95%CI, 30.5-66.1), which was similar to the progression in those with an EBA IA-2A PLS >4.0 (58.7% (95%CI, 37.1-72.8).

**Figure 1.**
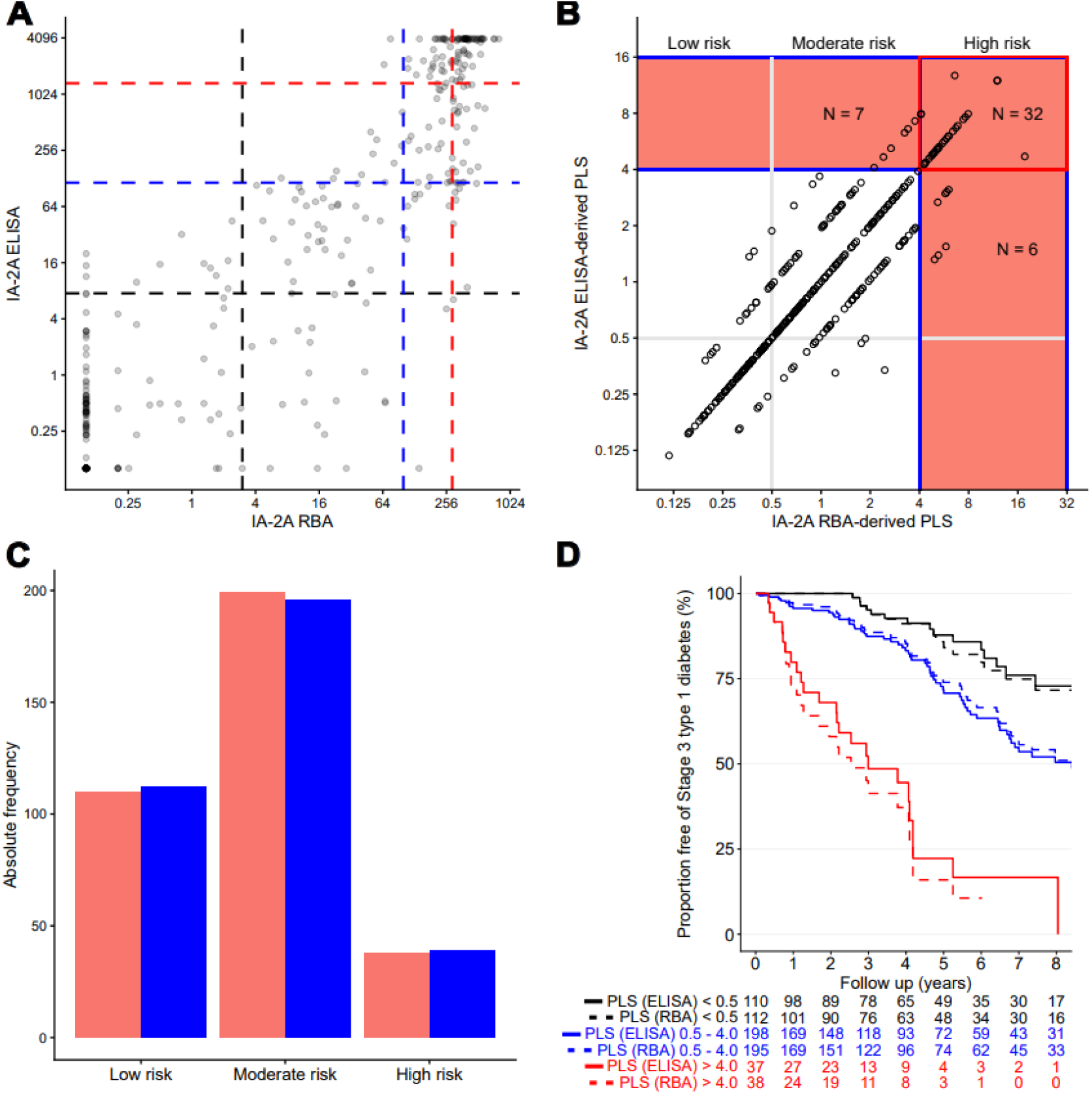
Comparison of IA-2A measured by ELISA and RBA in children with Stage 1 type 1 diabetes. A. Comparison of values obtained in the ELISA (y axis) and RBA (x axis) from 349 children with stage 1. The vertical dashed lines indicate the IA-2A thresholds in the RBA that define IA-2A categories used in calculating the progression likelihood score (PLS) and the horizontal dashed lines are the corresponding thresholds calculated for the ELISA. The darker dots indicate more than one data point. B. Comparison of PLS values derived from the ELISA (y axis) and RBA (x axis) IA-2A measurements in 347 children with stage 1. The low, moderate and high risk groups are indicated, which the high risk group (PLS >4.0) shaded in pink. The number of children with scores in the concordant and discordant high risk groups are indicated. C. Histogram indicating the number of children in the low, moderate and high risk PLS groups using the RBA IA-2A measurements (blue) or the ELISA IA-2A measurements (red). D. Stratification of progression from stage 1 to stage 3 type 1 diabetes by the PLS calculated using IA-2A units measured by RBA (dashed lines) and ELISA (solid lines). Children were categorized using previously defined PLS thresholds as <0.5 (low, black line), 0.5 – 4.0 (intermediate, blue line) and >4.0 (high, red line). Progression differed significantly among categories in both the PLS derived from the ELISA IA-2A units (p<0.0001) and the RBA IA-2A units (p<0.0001). The numbers underneath the x axis indicate the number remaining at each year of follow-up.

## Conclusions

The PLS relies on IA-2A titre categories. Here, we show that although there are some discrepancies between the historical harmonized radiobinding assay and the certified commercial ELISA for IA-2A measurement, a PLS >4.0 using either assay in children with stage 1 type 1 diabetes was associated with a faster rate of progression to stage 3 disease approaching the rates seen in children with stage 2 disease^2^. A similar finding was previously reported when the MSD ECL assay was used for IA-2A measurement^4^. These findings demonstrate that the PLS robustly identifies a subgroup of children with stage 1 disease who are at high risk of rapid progression to stage 3 and support its use to select individuals for clinical trials, irrespective of the IA-2A assay employed.

## Acknowledgments

This study was supported by a grant from the EASD-Novo Nordisk Foundation Diabetes Prize for Excellence (NNF22SA0081044), and the German Center for Diabetes Research ([DZD] e.V.).

## Ethics statement

The Fr1da study (was approved by the Ethical Review Board (Technical University Munich; protocol #808040014).

Study Registration: NCT04039945

## Data availability

All data produced in the present study are available upon reasonable request to the authors.

## Author Contributions

Design EB, AZ

Conduct/Data collection MS

Analysis EB, AW

Writing manuscript EB, AZ

## References

(1) Insel, R.A.; Dunne, J.L.; Atkinson, M. A.; Chiang, J. L.; Dabelea, D.; Gottlieb, P. A.; Greenbaum, C. J.; Herold, K. C.; Krischer, J. P.; Lernmark, Å.; et al. Staging Presymptomatic Type 1 Diabetes: A Scientific Statement of JDRF, the Endocrine Society, and the American Diabetes Association. Diabetes Care 2015, 38 (10), 1964–1974. DOI: 10.2337/dc15-1419 (accessed 1/21/2026).

(2) Hummel, S.; Koeger, M.; Bonifacio, E.; Ziegler, A.-G. Dysglycaemia definitions and progression to clinical type 1 diabetes in children with multiple islet autoantibodies. The Lancet Diabetes & Endocrinology 2025, 13 (1), 10–12. DOI: 10.1016/S2213-8587(24)00337-1 (accessed 2026/01/20).

(3) Weiss, A.; Zapardiel-Gonzalo, J.; Voss, F.; Jolink, M.; Stock, J.; Haupt, F.; Kick, K.; Welzhofer, T.; Heublein, A.; Winkler, C.; et al. Progression likelihood score identifies substages of presymptomatic type 1 diabetes in childhood public health screening. Diabetologia 2022, 65 (12), 2121–2131. DOI: 10.1007/s00125-022-05780-9 From NLM.

(4) Weiss, A.; Chakievska, L.; Achenbach, P.; Hergl, M.; Hummel, S.; Ott, R.; Scholz, M.; Winkler, C.; Bonifacio, E.; Ziegler, A.-G. Stratifying the Rate of Disease Progression by Progression Likelihood Scores in Children and Adolescents With Stage 1 and Stage 2 Type 1 Diabetes in Germany. Diabetes Care 2025, 49 (2), 352–360. DOI: 10.2337/dc25-2184 (accessed 1/21/2026).

(5) Achenbach, P.; Warncke, K.; Reiter, J.; Naserke, H. E.; Williams, A. J.; Bingley, P. J.; Bonifacio, E.; Ziegler, A. G. Stratification of type 1 diabetes risk on the basis of islet autoantibody characteristics. Diabetes 2004, 53 (2), 384–392. DOI: 10.2337/diabetes.53.2.384 From NLM.

(6) Bonifacio, E.; Yu, L.; Williams, A. K.; Eisenbarth, G. S.; Bingley, P. J.; Marcovina, S. M.; Adler, K.; Ziegler, A. G.; Mueller, P. W.; Schatz, D. A.; et al. Harmonization of glutamic acid decarboxylase and islet antigen-2 autoantibody assays for national institute of diabetes and digestive and kidney diseases consortia. J Clin Endocrinol Metab 2010, 95 (7), 3360–3367. DOI: 10.1210/jc.2010-0293 From NLM.

(7) Ziegler, A.-G.; Kick, K.; Bonifacio, E.; Haupt, F.; Hippich, M.; Dunstheimer, D.; Lang, M.; Laub, O.; Warncke, K.; Lange, K.; et al. Yield of a Public Health Screening of Children for Islet Autoantibodies in Bavaria, Germany. JAMA 2020, 323 (4), 339–351. DOI: 10.1001/jama.2019.21565 (accessed 1/21/2026).

(8) Committee, A. D. A. P. P. 2. Diagnosis and Classification of Diabetes: Standards of Care in Diabetes—2025. Diabetes Care 2024, 48 (Supplement_1), S27–S49. DOI: 10.2337/dc25-S002 (accessed 1/21/2026).

